# Wearable-derived physiological features for trans-diagnostic disease comparison and classification in the All of Us longitudinal real-world dataset

**DOI:** 10.64898/2026.04.07.26350352

**Authors:** Xueyan Huang, Chihcheng Hsieh, Quan Nguyen, Miguel E. Rentería, Puya Gharahkhani

**Author notes:** These authors contributed equally to this work (Miguel E. Rentería, Puya Gharahkhani).

## Abstract

Wearable-derived physiological features have been associated with disease risk, but most current studies focus on single conditions, limiting understanding of cross-disease patterns. This study adopts a trans-diagnostic approach to examine whether wearable data capture shared and condition-specific physiological signatures across multiple chronic conditions spanning physical and mental health, and then evaluates the utility of these features for disease classification. A total of 9,301 patients with at least 21 days of consecutive FitBit data from the All of Us Controlled Tier Dataset version 8 were analyzed. Disease subcohorts included cardiovascular disease (CVD), diabetes, obstructive sleep apnea (OSA), major depressive disorder (MDD), anxiety, bipolar disorder, and attention-deficit/ hyperactivity disorder (ADHD), chosen based on prevalence and relevance. Logistic regression and XGBoost models were fitted for each disease subcohort versus the control cohort. We found that compared to using just baseline demographic and lifestyle features, incorporating wearable-derived features enabled improved classification performance in all subcohorts for both models, except for ADHD where improvement was mainly observed for ROC-AUC in logistic regression model likely due to the smaller sample size in ADHD subcohort. The largest performance gains were observed in MDD (increase in ROC-AUC of 0.077 for Logistic regression, 0.071 for XGBoost; p *<* 0.001) and anxiety (increase in ROC-AUC of 0.077 for logistic regression, 0.108 for XGBoost; p *<* 0.001). This study provides one of the first comprehensive transdiagnostic evaluations of wearable-derived features for disease classification, highlighting their potential to enhance risk stratification in the real-world setting as a practical complement to clinical assessments and providing a foundation to explore more fine-grained wearable data.

**Author summary:** Wearable devices such as fitness trackers and smartwatches are becoming increasingly popular and affordable, providing continuous measurements of heart rate, physical activity, and sleep. Alongside the growing digitization of health records, this creates new opportunities for large-scale, real-world health studies. In this study, we analyzed wearable-derived physiological patterns across a range of chronic conditions spanning both physical and mental health to better understand how these signals relate to disease risk. We found that incorporating wearable-derived heart rate, activity and sleep features improved disease risk classification across several conditions, with particularly strong gains for major depressive disorder and anxiety. By examining how individual features contributed to model predictions, we also identified meaningful associations between physiological signals and disease risk. For example, both duration and day-to-day variation of deep and rapid eye movement (REM) sleep were associated with increased risk in certain conditions. Our study supports the development of real-time, automated tools to assess disease risk alongside clinical care.

## Introduction

The use of physiological data collected from non-invasive wearable devices has grown rapidly in recent years, driven by advances in sensor technology and machine learning methods that enable continuous, real-world health monitoring [1]. Wearable-derived metrics such as heart rate, physical activity and sleep have shown promise in tracking disease progression and predicting adverse events across a range of chronic conditions including both physical and mental health conditions such as cardiovascular conditions [2, 3], depression, bipolar disorder [4], and anxiety [5]. However, many current studies on machine learning-based disease classification focus on single conditions under controlled conditions and may not reflect the complexity of real-world clinical presentations, which are often heterogeneous and multimorbidity is common. While there are studies that take a trans-diagnostic approach to disease comparison [6], few have explicitly explored and quantified the utility of wearable-derived features in classifying a broad array of chronic health conditions in a real world dataset.

To address this gap, this study aims to: (1) characterize the differences and similarities in physiological patterns across multiple common chronic health conditions, spanning both physical and mental health; and subsequently (2) evaluate the predictive utility of wearable-derived features for disease classification in a multi-diagnosis setting. This study combines inferential statistics and predictive modeling to inform more comprehensive future modeling efforts.

## Materials and Methods

This study uses the All of Us Controlled Tier Dataset version 8 (v8), a longitudinal health dataset from participants across the United States collected by the National Institutes of Health (NIH). Compared to previous versions, v8 is the first version to include Wearables Enhancing All of Us Research (WEAR) initiative [7], increasing the sample size of patients with both FitBit and EHR data from 14,908 in v7 [8] to 36,614 in v8. The FitBit data consists of 57% Bring-Your-Own-Device (BYOD) participants and 43% Wearables Enhancing All of Us Research (WEAR) participants, whom are provided with either a FitBit Charge 4™ tracker or FitBit Versa 3™ smartwatch. [7] [9] FitBit data collected include heart rate (HR), physical activity (PA) and sleep, both on a minute-level basis and a daily summary level.

Electronic health records (EHR) data was shared on a voluntary basis and standardized with Observational Medical Outcomes Partnership (OMOP) Common Data Model (CDM), which includes data harmonization, cleaning, and methods to de-identify patients such as removing explicit personal identifiers, generalizing date of birth. The privacy rules are summarized here: [10].

### Inclusion, exclusion criteria and cohort definition

Patients with at least 21 days of consecutive FitBit data and shared EHR records were included in the cohort. Daily summary level FitBit data were used for analysis; days were considered valid when FitBit summary statistics are available. In addition, patients must be at least 18 years old on the first day of 21 days of consecutive FitBit data. Patients missing key demographic variables such as age, biological sex and BMI were excluded.

We focused on seven disease subcohorts: cardiovascular disease (CVD), diabetes, obstructive sleep apnea (OSA), major depressive disorder (MDD), anxiety, bipolar disorder, and attention-deficit/ hyperactivity disorder (ADHD). These conditions were chosen as they are prevalent, clinically important, and known to impact physical activity, heart rate, and sleep patterns. Patients were assigned to one of seven disease subcohorts based on ICD-9 and ICD-10 codes. To map ICD-9 and ICD-10 codes into subcohorts, mappings from the R PheWAS analysis package were used to first map ICD-9 and ICD-10 codes into descriptions [11], which were then grouped into subcohorts according to the mapping in S1 Table. Participants meeting criteria for multiple diagnoses were assigned to multiple subcohorts. Patients without any of the target diagnoses, plus additional exclusion of major conditions based on ICD-9 and ICD-10 codes, were assigned to the control cohort. The full list of ICD-9 and ICD-10 codes excluded in control cohort is included in the S1 Table.

Furthermore, to ensure that diagnosis reflects current condition, the most recent diagnosis must be recorded no more than a maximum number of years before monitoring period as defined in sub-section Variables. Maximum number of years allowed between diagnosis and monitoring period varies according to subcohorts, as some diagnoses represent lifetime risk factors such as CVD, Type II Diabetes, and Bipolar while others, such as MDD and anxiety, can often go into full remission. The exact maximum number of years allowed were determined by balancing sample size with typical timelines for recurrence and persistence of each condition, as detailed below for each subcohort in Table 1.

**Table 1.** Maximum number of years allowed for most recent diagnosis before FitBit monitoring period.

| Maximum number of years allowed | Conditions |
| --- | --- |
| 10 years | CVD, Type II Diabetes |
| 5 years | Bipolar disorder |
| 3 years | OSA, ADHD |
| 2 years | MDD, Anxiety |

### Variables

A comprehensive set of demographic, lifestyle, and wearable-derived variables was included for cohort characterization and descriptive analyses. Demographic variables comprised age, sex, race, and body mass index (BMI). Lifestyle variables include alcohol consumption and smoking behavior. Wearable-derived variables included daily summary statistics of heart rate (HR), physical activity, and sleep calculated by FitBit using proprietary algorithms, which integrated raw sensor signals from 3-axis accelerometry and photoplethysmography (PPG). These summaries include FitBit internal data pre-processing and quality control procedures, and no additional data cleaning was performed. We defined a participant-specific **monitoring period** of 21 days during which the participant had valid FitBit summary statistics each day. Hereafter, we refer to this interval as the monitoring period. To capture both typical behavior and day-to-day variability, wearable measures were summarized as mean and standard deviation (STD) across the derived 21-day monitoring period for each participant. To capture finer daily dynamics, root mean squares of successive differences (RMSSD) of sleep features were also included. An interaction term between heart rate and activity was also included based on results from exploratory data analysis (EDA). Additional data processing, for example, further binning of categorical variables, was performed for subsequent modeling analyses. Smoking behavior was aggregated based on both lifetime and current smoking status and classified into the following categories: Never Smoker, Former Smoker, Current Smoker (some days), Current Smoker (every day), and Unknown for modeling. Descriptions of all variables compared descriptively are detailed in Table 2.

**Table 2.**
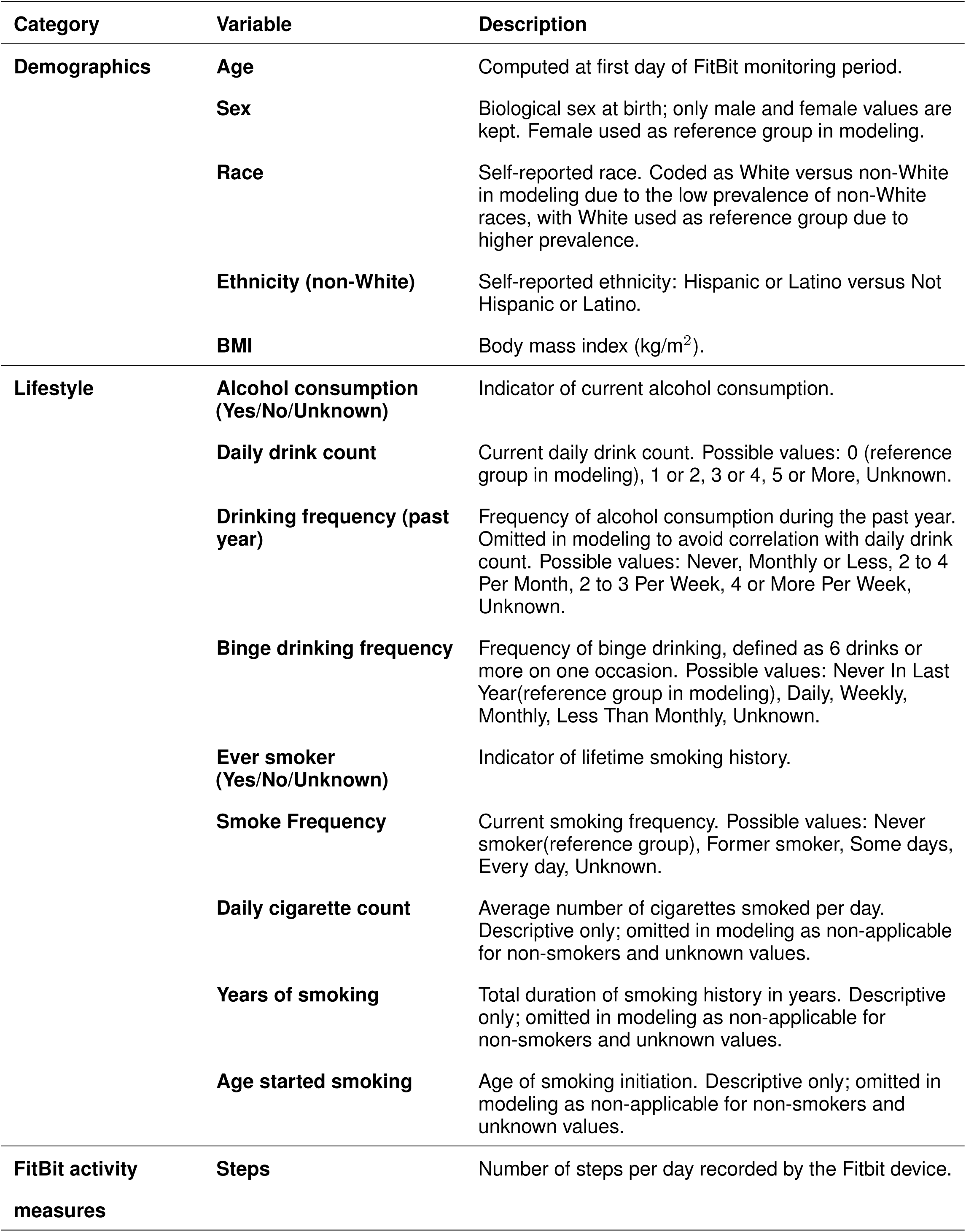
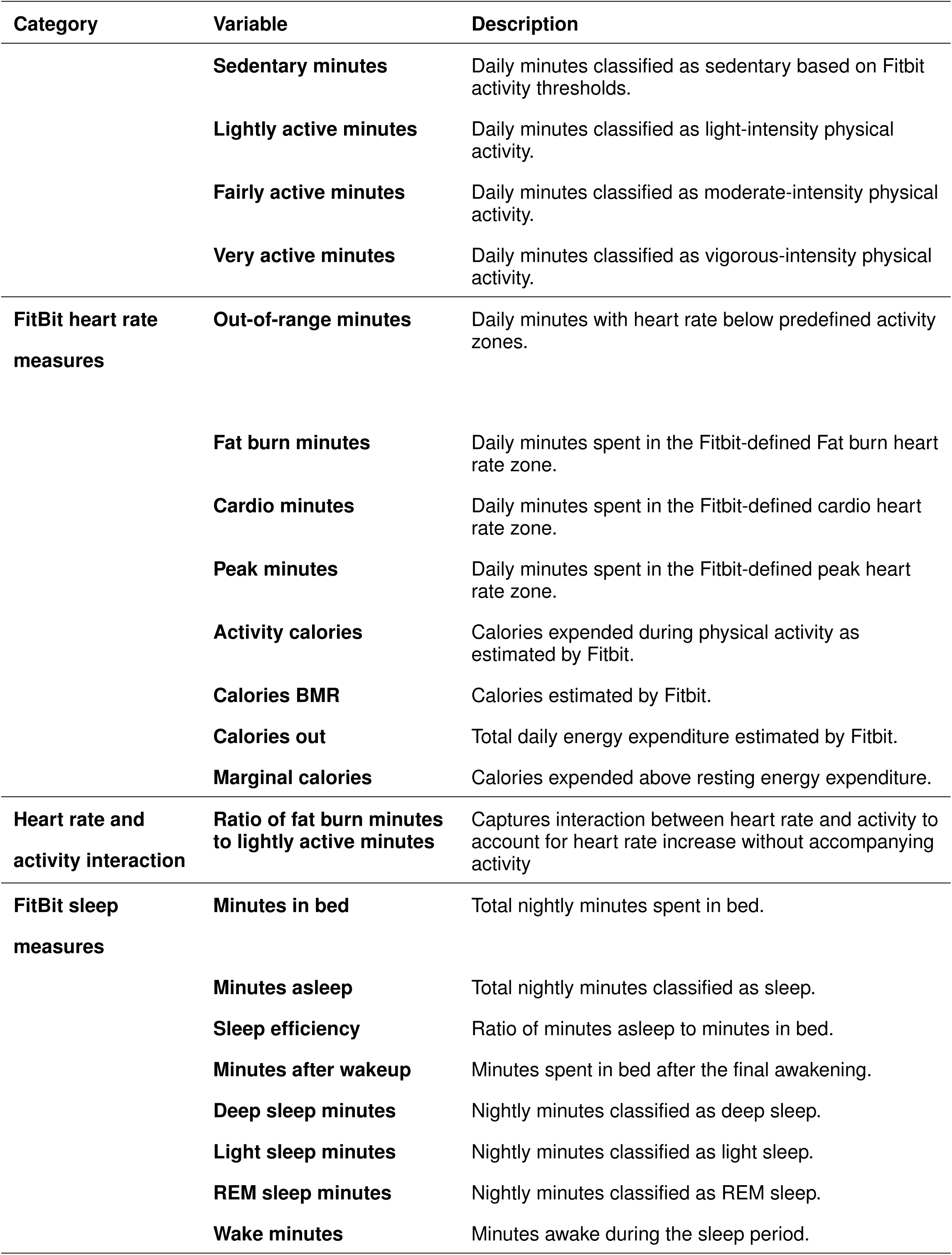
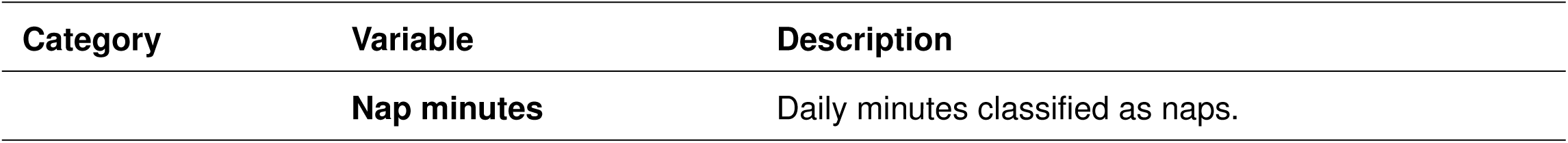
Description of demographic, lifestyle, and wearable-derived variables.

**Table 3.**
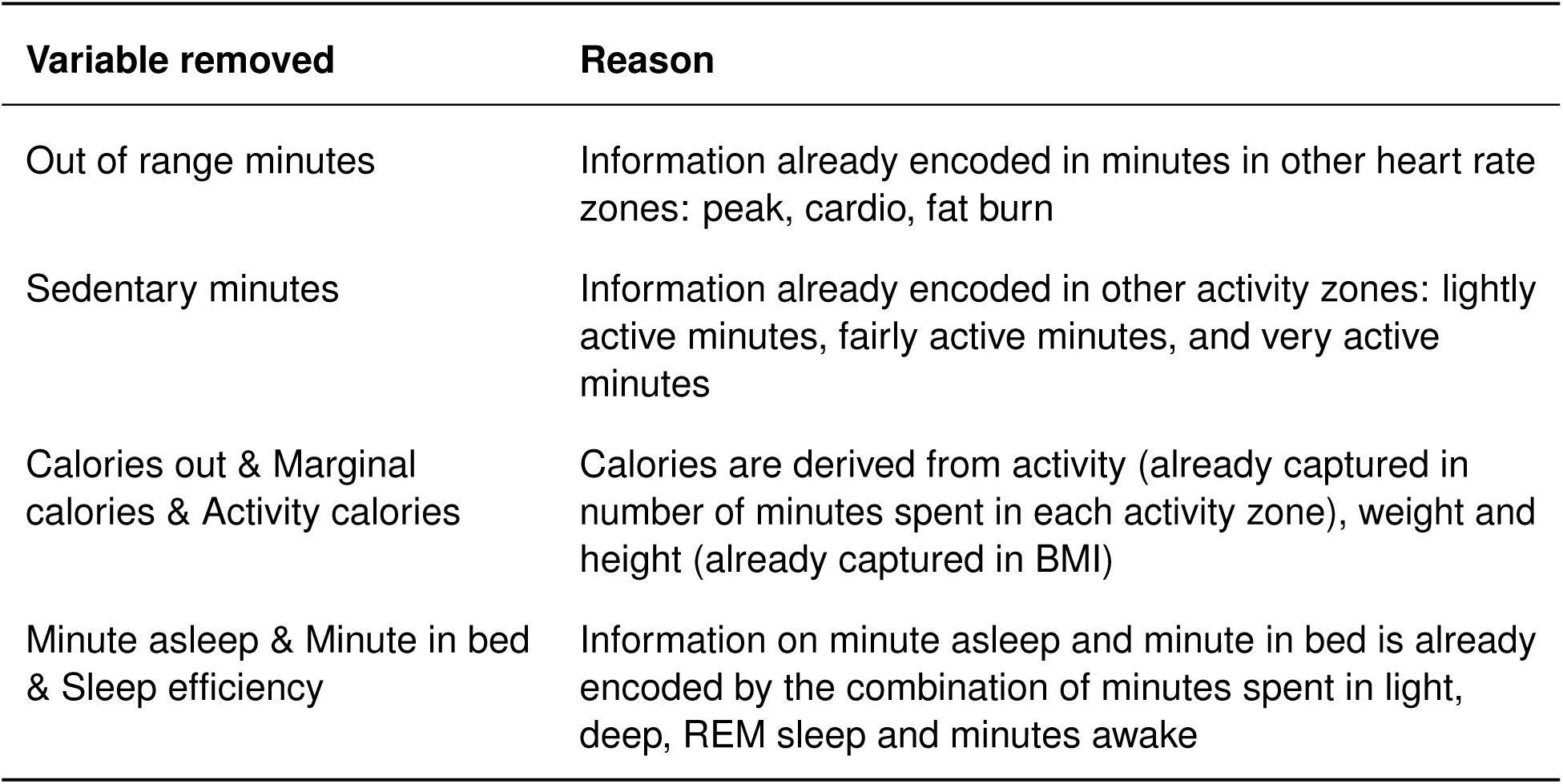
Wearable-derived features removed.

To limit multi-collinearity for modeling and to preserve the interpretaility of regression coefficients, feature selection on wearable-derived features was performed using a combination of domain knowledge and correlation analyses to obtain a more compact set of features that still capture essential heart rate, physical activity and sleep patterns. Pairwise Pearson correlation coefficients (*r*) among covariates were computed within the training dataset to identify highly correlated variables. Variance inflation factors (VIF) were then calculated to assess multicollinearity in regression-based models. The feature selection process was iterated until all VIFs are within an acceptable level. Variables with high statistical correlation but distinct interpretation were not removed. For example, high statistical correlations were identified between coefficient of variation for fairly active minute and very active minute (*r* = 0.81), mean and coefficient of variation for REM sleep (*r* =-0.73), mean and coefficient of variation for deep sleep (*r* =-0.76), and mean and coefficient of variation for nap minutes (*r* =-0.71), but all of these variables were kept in the model due to different interpretations. Data-driven feature selection using correlation and multi-collinearity analysis was conducted on training dataset only to prevent data leakage and optimistic bias in test set performance evaluation. The variables removed during feature selection process are detailed in Table **??**

## Statistical Analysis

Due to the nature of multi-morbidity, each disease area was treated as a separate label and a binary classification model was built for each disease area versus the control cohort. Each disease subcohort and the control cohort were split into 70% train and 30% test set. Both logistic regression and XGBoost models were then fitted on the curated set of features for each disease area versus the control cohort. During exploratory data analysis, potential non-linear effects and differences in model coefficients were identified, therefore including both models allowed us to capture potential non-linear relationships and to compare differences in feature effects.

For logistic regression models, to preserve the interpretability of regression coefficients, no regularization technique was used. P-values were adjusted for multiple testing using Benjamini-Hochberg procedure, a method to control Type I error in multiple comparisons by controlling the False Discovery Rate (FDR). For XGBoost models, hyperparameter tuning was performed using grid search with standard 5-fold cross-validation (GridSearchCV) on the training dataset. A grid of key hyperparameters including tree depth and number of estimators was systematically evaluated, and the combination of hyperparameters that achieved the best mean cross-validation classification accuracy was selected and used to train the final model. Feature importances for XGBoost models were evaluated using mean absolute SHAP values which represent the average magnitude of each feature’s contribution to the model’s predictions, providing an interpretable measure of feature influence across the dataset.

As XGBoost models are less sensitive towards multicollinearity than linear models, versions of XGBoost models fitted without feature selection and instead with full set of features were also evaluated. These models achieved similar performance as versions with feature selection, and hence feature selection was retained in XGBoost model to improve the robustness of feature importance values obtained in XGBoost models.

### Class imbalance

The class imbalance problem was moderate, ranging from the lowest event rate in ADHD subcohort (14%) and Bipolar subcohort (17%) to close to 38-46% in all the other subcohorts.

To address the lower event rate of ADHD and Bipolar, both unweighted models and class-weighted models were evaluated in these two subcohorts for both logistic regression and XGBoost. Model performance was assessed using discrimination (ROC-AUC score) and calibration (Brier score and Brier Skill Score). Although weighting modestly increased recall when classification threshold was set to 0.5 without optimization, ROC-AUC score was comparable between approaches, whereas calibration deteriorated substantially under weighting; negative Brier Skill Scores observed in logistic regression models indicated inferior performance to the reference prevalence model. Therefore, we retained the unweighted models to preserve probability calibration and instead calculated accuracy, precision, recall, and F1 metrics using adjusted decision thresholds that maximises F1 score in the training dataset across all subcohorts. The same thresholds were then applied to test dataset to calculate accuracy, precision, recall, and F1 scores.

### Performance metrics

Each dataset was split into 70% for training and 30% for testing. Models training and related tasks such as feature selection and optimizing classification threshold were conducted on the training dataset only to prevent data leakage; model performance was evaluated using the unseen test dataset across multiple metrics. **Discrimination**, or the model’s ability to distinguish between cases and non-cases, was assessed using accuracy, precision, recall, F1 score, and the area under the receiver operating characteristic curve (ROC-AUC Score). **Calibration** was assessed using the calibration curves, providing visual assessments of agreement between predicted and actual risks by comparing predicted probabilities with observed outcome frequencies across quantiles of predicted risk. Perfect calibration is indicated by alignment with the diagonal line. Overall calibration accuracy was quantified using the Brier score [12], which measures the mean squared difference between predicted probabilities and observed outcomes, with lower values indicating better calibrated predictions. For easier interpretation, the Brier Skill Score (BSS) was then calculated as below [13]:

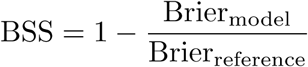

which expresses the proportional improvement in Brier score compared with a reference model, defined in this study as a model that always predicts disease prevalence. BSS values greater than zero indicate improved predictive performance over the reference model, while values less than or equal to zero indicate no improvement or worse performance. All performance metrics were then evaluated for robustness using confidence intervals derived from 1,000 bootstrap samples. ROC-AUC and BSS were considered to be core performance metrics measuring discrimination and calibration respectively, among others. To assess whether incorporating wearable-derived features improved model performance, base models including only demographic and lifestyle features were fitted using the same training dataset and hyperparameters, and subsequently compared with models that also included wearable-derived features on the same held-out test dataset. Differences in model performance between base models and models incorporating FitBit features were evaluated using two-sided hypothesis tests. Specifically, differences in ROC-AUC were assessed using the DeLong test, whereas 1,000-resample paired bootstrapping was applied for all other metrics.

### Medication and sensitivity analysis

Certain medications are known to influence sleep patterns and heart rate, and some may indicate suspected conditions even in the absence of a formal diagnosis. Therefore, a stricter definition of the control cohort with an additional exclusion criterion to exclude patients with prescriptions that could affect sleep or heart rate was investigated as a potential alternative to the control cohort with only the inclusion and exclusion criteria described in sub-section Inclusion, exclusion criteria and cohort definition. This cohort is hereafter referred to as the **’Clean Control cohort’**, whereas the version of control cohort without any restrictions on medication is referred to as the **’Control Cohort’**. The full list of medications excluded from Clean Control Cohort are included in S2 Table. Control Cohort was compared with the Clean Control cohort on all demographic, lifestyle and wearable-derived features to determine whether the distribution of covariates differ significantly among the two versions of control cohort. Continuous variables were compared using Welch’s t-test, with effect sizes reported as Cohen’s d to indicate the standardized mean differences. Approximate normality was assumed based on large sample size.

Categorical variables were compared using Chi-square or Fisher’s exact tests as appropriate, with Cramér’s V used to quantify effect sizes. All tests were two-sided, with p-values smaller than 0.05 considered statistically significant. The full comparison of Control Cohort vs Clean Control Cohort are included in S4 Table. Control Cohort was used in the final modeling due to similar covariate distribution to Clean Control Cohort and larger sample size.

## Results

### Cohort attrition and descriptives

A total of 9,301 people were included in the descriptive and modeling analyses after inclusion and exclusion criteria in Table 4 were applied, with subcohort sizes: Control (N = 2,830), CVD (N = 2,289), Diabetes (N = 1,783), OSA (N = 2,124), MDD (N = 1,867), anxiety (N = 2,362), Bipolar (N = 561), ADHD (N = 471). The version of control cohort defined without further medication exclusion was used due to the larger sample size as elaborated in Materials and Methods section, sub-section Medication and sensitivity analysis.

**Table 4.** Cohort attrition.

| Inclusion/ Exclusion criteria | N |
| --- | --- |
| Patients who provided FitBit data | N = 52,334 |
| Patients who provided EDHR data | N = 36,614 |
| Patients with at least 21 days of FitBit data | N = 16,519 |
| At least 18 years old at the time of FitBit data collection | N = 16,517 |
| Assigned to one of disease subcohorts or control | N = 11,038 |
| Diagnoses filtered based on recency criteria | N = 9,872 |
| Key demographic variables available | N = 9,301 |

Key demographic and lifestyle features for each subcohort are described in Table 5. Median age of the subcohorts ranged from 39 years old in ADHD subcohort (youngest) and 68 years old in CVD subcohort (oldest), majority Female with CVD, Diabetes and OSA having a larger proportion of males compared to the rest. The subcohorts were also predominantly White. Median BMI was 26.2 for control cohort and ranged from 28.9 in ADHD subcohort (lowest) to 32.1 in OSA subcohort (highest). Majority of the cohorts reported alcohol consumption while smoking habits vary, with control cohort having 28.7% reporting ever smoking (100 cigarettes lifetime), while the rest of the subcohorts ranged from 34.8% in ADHD subcohort (lowest) and 47.2% in CVD subcohort (highest). Full descriptives of demographic, lifestyle and wearable-derived features are summarized in S3 Table.

**Table 5.** Descriptives of key demographic and lifestyle features across subcohorts.

|  |  |  | Control | CVD | Diabetes | OSA | MDD | Anxiety | Bipolar | ADHD |
| --- | --- | --- | --- | --- | --- | --- | --- | --- | --- | --- |
| **Age** | **Median** |  | 48.00 | 68.00 | 62.00 | 61.00 | 51.00 | 48.00 | 48.00 | 39.00 |
| **Sex at Birth** | **N (%)** | **Female** | 2055<br>(72.61) | 1202<br>(52.51) | 1081<br>(60.63) | 1224<br>(57.63) | 1445<br>(77.40) | 1855<br>(78.54) | 408<br>(72.73) | 346<br>(73.46) |
| **Race** | **N (%)** | **White** | 2104<br>(74.35) | 1961<br>(85.67) | 1343<br>(75.32) | 1707<br>(80.37) | 1510<br>(80.88) | 1955<br>(82.77) | 457<br>(81.46) | 398<br>(84.50) |
| **Ethnicity** | **N (%)** | **Hispanic or Latino** | 293<br>(10.35) | 77 (3.36) | 117 (6.56) | 116 (5.46) | 125 (6.70) | 155 (6.56) | 32 (5.70) | 27 (5.73) |
| **BMI** | **Median** |  | 26.22 | 28.50 | 31.80 | 32.10 | 29.90 | 29.10 | 30.60 | 28.90 |
| **Alcohol consumption** | **N (%)** | **Yes** | 2731<br>(96.50) | 2224<br>(97.16) | 1704<br>(95.57) | 2056<br>(96.80) | 1810<br>(96.95) | 2298<br>(97.29) | 542<br>(96.61) | 458<br>(97.24) |
| **Ever smoked** | **N (%)** | **Yes** | 812<br>(28.69) | 1081<br>(47.23) | 745<br>(41.78) | 851<br>(40.07) | 709<br>(37.98) | 868<br>(36.75) | 270<br>(48.13) | 164<br>(34.82) |

### Comparison of model performance

Key model performance metrics assessed using ROC-AUC score and Brier Skill Score (BSS) with 95% confidence intervals are summarized in Fig 1. Differences in core model performance metrics are summaried in Table 6. At 5% significance level, incorporating FitBit features significantly improved the AUC-ROC across all subcohorts except for the XGBoost model in the ADHD subcohort. Improvements in Brier Skill Score were also observed across all subcohorts, except for both models in the ADHD subcohort and the XGBoost model in the CVD subcohort. No statistically significant difference in model performance was detected between logistic regression with FitBit features and XGBoost models with FitBit features, as summaried in Table 7. Full model performance metrics with 95% confidence intervals and comparisons are detailed in S5 Table Both logistic regression and XGBoost models achieved good calibration in subcohorts CVD, diabetes, OSA, MDD and anxiety as shown in Figure 2, approximating reference perfect calibration curves. For bipolar and ADHD subcohorts, calibration curves aligned well with observed outcomes at lower predicted probabilities but deviated increasingly at higher predicted probabilities (*>* 0.6), indicating less reliable risk scores when predicted probabilities are high, possibly due to the smaller sample sizes in both of these subcohorts.

**Fig 1.**
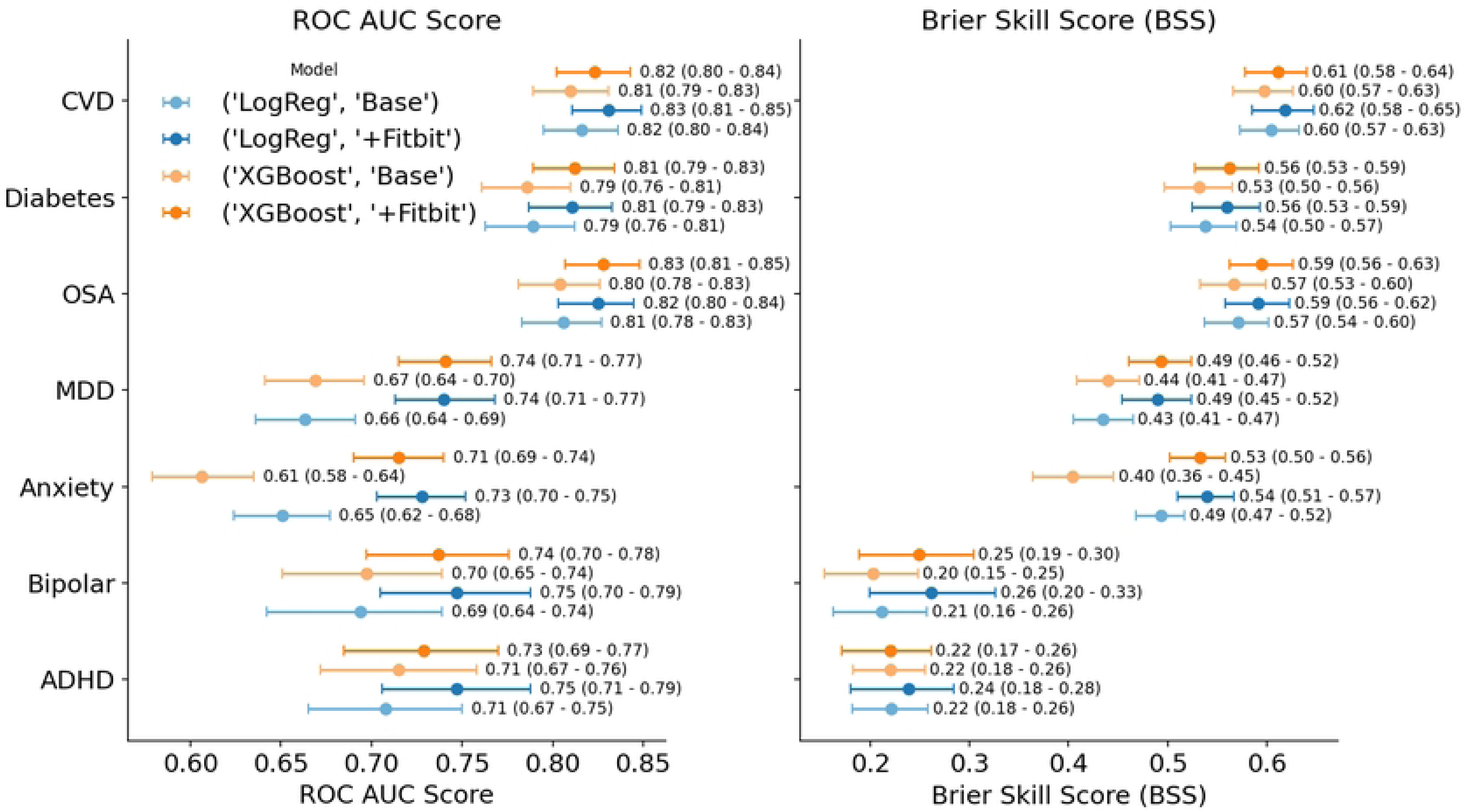
ROC-AUC Score and Brier Skill Score (BSS) across subcohorts and models A. ROC-AUC Score. B. Brier Skill Score (BSS).

**Fig 2.**
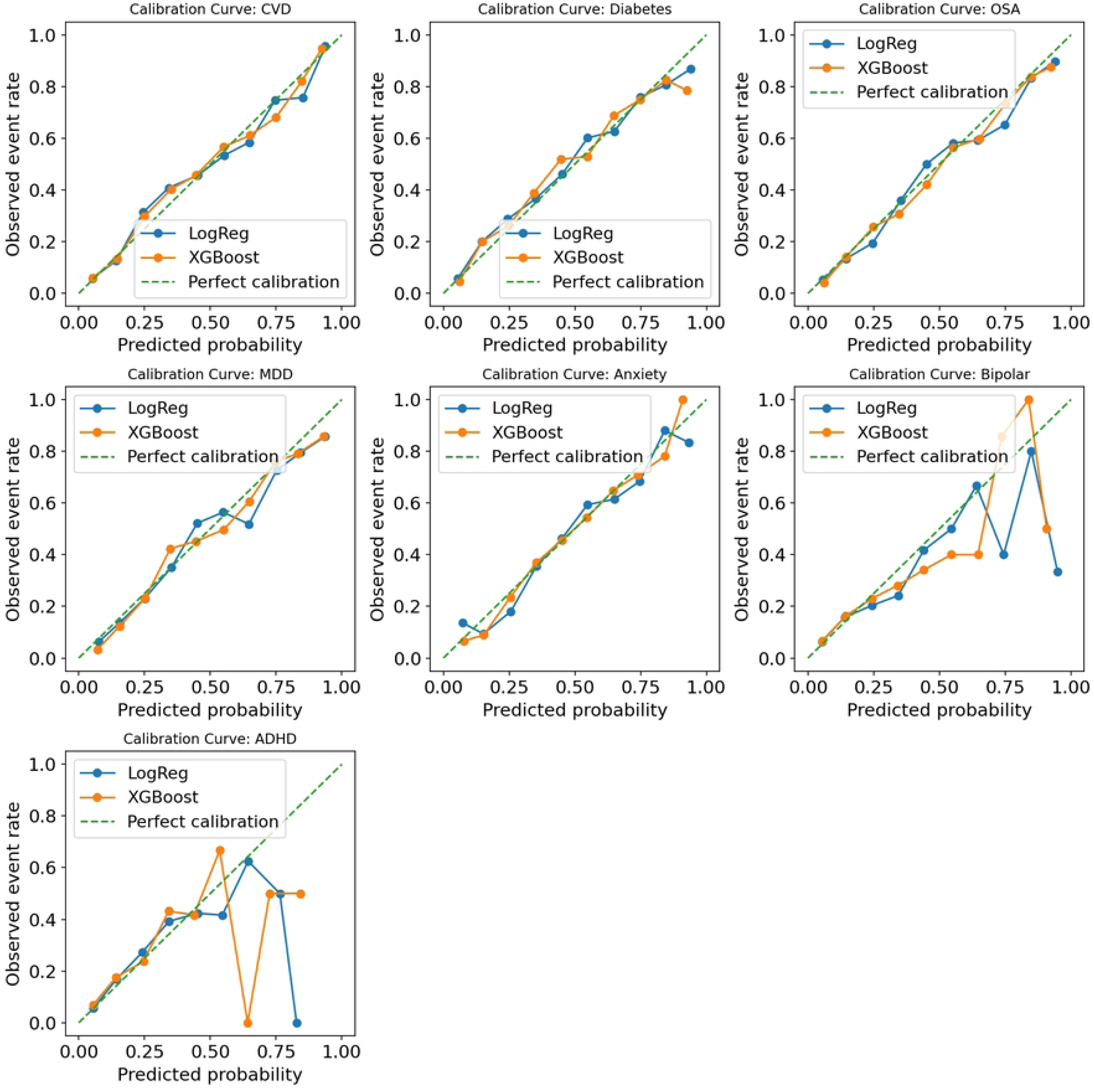
Calibration curves across subcohorts

**Table 6.** Improvement in Discrimination (AUROC) and Calibration (BSS) after incorporating FitBit features (95% CI)

|  |  | ROC AUC Score |  | Brier Skill Score (BSS) |  |
| --- | --- | --- | --- | --- | --- |
|  |  | Difference (95% CI) | p-value | Difference (95% CI) | p-value |
| **CVD** | **LogReg** | 0.015 (0.005 - 0.026) | 0.003* | 0.015 (0.001 - 0.027) | 0.032* |
|  | **XGBoost** | 0.013 (0.001 - 0.025) | 0.016* | 0.013 (-0.001 - 0.027) | 0.072 |
| **Diabetes** | **LogReg** | 0.022 (0.010 - 0.034) | < 0.001* | 0.022 (0.008 - 0.036) | 0.002* |
|  | **XGBoost** | 0.026 (0.011 - 0.040) | < 0.001* | 0.030 (0.013 - 0.048) | < 0.001* |
| **OSA** | **LogReg** | 0.019 (0.006 - 0.031) | 0.002* | 0.020 (0.006 - 0.035) | 0.006* |
|  | **XGBoost** | 0.024 (0.010 - 0.038) | < 0.001* | 0.028 (0.012 - 0.044) | < 0.001* |
| **MDD** | **LogReg** | 0.077 (0.052 - 0.102) | < 0.001* | 0.055 (0.035 - 0.074) | < 0.001* |
|  | **XGBoost** | 0.071 (0.045 - 0.098) | < 0.001* | 0.052 (0.032 - 0.071) | < 0.001* |
| **Anxiety** | **LogReg** | 0.077 (0.054 - 0.100) | < 0.001* | 0.048 (0.033 - 0.063) | < 0.001* |
|  | **XGBoost** | 0.108 (0.075 - 0.142) | < 0.001* | 0.129 (0.098 - 0.160) | < 0.001* |
| **Bipolar** | **LogReg** | 0.052 (0.013 - 0.092) | 0.005* | 0.049 (0.007 - 0.088) | 0.018* |
|  | **XGBoost** | 0.039 (-0.004 - 0.083) | 0.039* | 0.046 (0.002 - 0.092) | 0.038* |
| **ADHD** | **LogReg** | 0.040 (0.001 - 0.078) | 0.023* | 0.018 (-0.016 - 0.050) | 0.296 |
|  | **XGBoost** | 0.012 (-0.036 - 0.060) | 0.312 | 0.000 (-0.040 - 0.039) | 0.972 |

**Table 7.** Difference in performance between XGBoost and Logistic Regression model (95% CI)

|  |  | ROC AUC Score |  | Brier Skill Score (BSS) |  |
| --- | --- | --- | --- | --- | --- |
|  |  | Difference (95% CI) | p-value | Difference (95% CI) | p-value |
| **CVD** |  | -0.008 (-0.016 - 0.001) | 0.959 | -0.008 (-0.018 - 0.003) | 0.160 |
| **Diabetes** |  | 0.001 (-0.010 - 0.013) | 0.401 | 0.002 (-0.013 - 0.016) | 0.806 |
| **OSA** |  | 0.003 (-0.007 - 0.013) | 0.277 | 0.004 (-0.009 - 0.017) | 0.572 |
| **MDD** |  | 0.000 (-0.015 - 0.015) | 0.482 | 0.003 (-0.010 - 0.015) | 0.686 |
| **Anxiety** |  | -0.013 (-0.028 - 0.002) | 0.958 | -0.008 (-0.019 - 0.002) | 0.140 |
| **Bipolar** |  | -0.010 (-0.037 - 0.017) | 0.772 | -0.012 (-0.052 - 0.026) | 0.500 |
| **ADHD** |  | -0.020 (-0.054 - 0.015) | 0.868 | -0.018 (-0.050 - 0.013) | 0.264 |

**Table 8.**
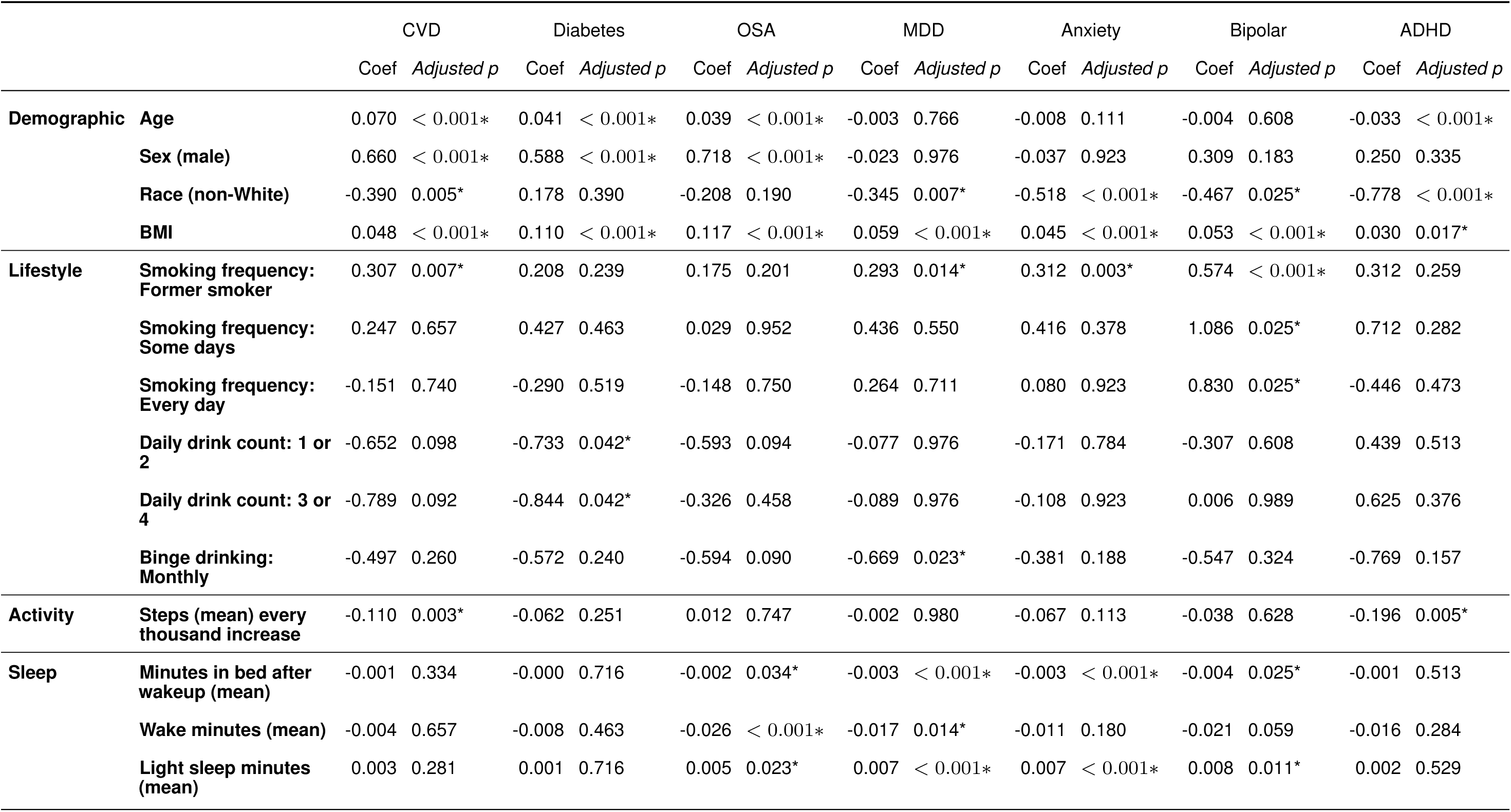
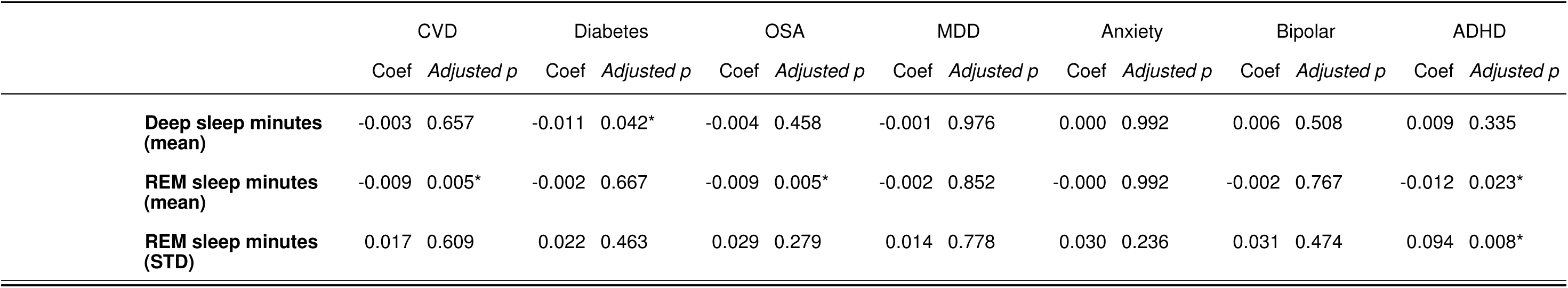
Regression coefficients across subcohorts at 5% significance level with Benjamini–Hochberg (BH) adjusted p-values.

### Logistic regression coefficients and XGBoost feature importance

For logistic regression models incorporating full wearable-derived features, variables with an adjusted statistical significance of 5% in at least one of the disease subcohorts are presented and compared below in Table 8, with the full regression coefficients for each subcohorts included in S6 Table. Feature importances for XGBoost models calculated as mean absolute SHAP values on the test dataset are included in S7 Table.

### Demographic features

Older age is associated with higher risk in CVD, Diabetes and OSA and lower risk in ADHD. Biological male sex is associated with higher disease risk in CVD, Diabetes, and OSA. Race non-White is associated with a lower disease risk in CVD, MDD, anxiety, Bipolar, and ADHD. A higher BMI is consistently associated with increased disease risk in all subcohorts.

### Lifestyle features

Former smoking is associated with increased disease risk in CVD, MDD, anxiety, and Bipolar, whereas current smoking (some days and every day) is associated with increased disease risk in Bipolar subcohort. Daily drink count of 1 or 2 and 3 or 4 (compared to 0 daily drink count) are associated with lower disease risk in Diabetes, and a monthly frequency of binge drinking (defined as 6 drinks or more) compared to no binge drinking at all, is associated with a lower disease risk in MDD.

### Wearable-derived features

For activity measures, higher number of steps is associated with lower disease risk in CVD and ADHD. For sleep features, minutes spent in bed after wake up is associated with lower disease risk in CVD, MDD, anxiety, and Bipolar. Higher mean light sleep minutes is associated with higher disease risk in OSA, MDD, anxiety, and Bipolar. Higher mean deep sleep minutes is associated with lower disease risk in Diabetes while higher mean REM sleep minutes is associated with lower disease risk in CVD, OSA and ADHD. Higher variation in REM sleep minutes is associated with higher disease risks in ADHD.

## Discussion

### Improvement in model performance from wearable-derived features

Incorporating FitBit wearable-derived features improved model performance across subcohorts, with the improvement in ROC-AUC most pronounced in MDD (from 0.67 to 0.74 for XGBoost, from 0.66 to 0.74 for Logistic regression) and anxiety (0.61 to 0.71 for XGboost, 0.65 to 0.73 for logistic regression) subcohorts. The improvements in classification of physical health conditions, namely CVD, Diabetes, and OSA are statistically significant but of smaller magnitudes, with the marginal increase in ROC-AUC in the 0.01 - 0.03 range. A smaller improvement in model performance in physical conditions could be because these conditions are already strongly predicted by demographic and lifestyle characteristics, as evidenced by the high ROC-AUC scores (around 0.8 in base models). MDD and anxiety are closely related conditions with high comorbidity rate and shared symptoms such as sleep disturbance, restlessness, fatigue etc; the symptoms are often episodic in nature, which might be a reason why wearable-derived features are more effective at distinguishing these conditions than just baseline demographic and lifestyle factors.

Moderate performance improvements were also observed in Bipolar subcohort (ROC-AUC from 0.7 to 0.74 for XGBoost, from 0.69 to 0.75 for logistic regression) and ADHD subcohort (ROC-AUC from 0.71 to 0.75 for logistic regression). However, while models in Bipolar and ADHD subcohorts have good discrimination, calibration lags behind compared to the other four subcohorts, with BSS around 0.25 as compared to the other five subcohorts with BSS ranging from 0.4 - 0.6. In the calibration curves presented in Figure 2. It can be observed that for these two subcohorts, predicted probabilities from 0 to 0.6 roughly approximate observed event rate, and at predicted probabilities above 0.6, predicted probabilities start to deviate from observed event rate, indicating less reliable risk scores at high predicted probabilities. A possible reason is smaller sample size in these two subcohorts and small high-risk subgroups, leading to less stable learning and less reliable predictions at higher risk. While there are techniques to calibrate model predictions such as regularization, such techniques were not employed to preserve the interpretability of logistic regression model coefficients.

### Interpretation of regression coefficients and feature importance

These associations should be interpreted with caution, as regression coefficients reflect conditional relationships within the multivariable framework rather than robust independent effects. Moreover, the observed associations have not been validated in external cohorts and may therefore reflect characteristics specific to the current cohort rather than the broader population. Consequently, regression coefficients and reported feature importance should not be interpreted causally, but rather as guides for further, more detailed investigations.

### Alcohol intake and smoking habit

A somewhat surprising result is that a monthly frequency of binge drinking is associated with decreased disease risk in MDD, and also that compared to non-drinkers, a daily drinking count of 1-4 is associated with decreased disease risk in Diabetes, as detailed in Results section, sub-section Lifestyle features. While some non-randomised epidemiological studies have found statistically significant association between moderate alcohol consumption and chronic disease risk, such effects are possibly due to reverse causation, for example, reduced alcohol intake after diagnosis, or residual confounding with variables such socioeconomic status, diet that have not been accounted for in the analysis, rather than a true protective effect. It should, however, be noted that non-drinkers makes up a small percentage of the cohort (3-4%) and that the regression coefficients for alcohol drinking were observed to be somewhat unstable with changes in covariate combinations, indicating that these coefficients cannot be interpreted as independent effects due to possible co-linearity and unobserved confounders such as socioeconomic status, diet, etc.

Another noteworthy result is that former smoking is still associated with higher disease risk in CVD, MDD, Axneity and Bipolar despite current non-smoker status. This is possibly due to residual effects of prior smoking, and potentially, smoking cessation following disease onset. However, as smoking status was assessed at the survey date rather than at diagnosis, the temporal relationship cannot be definitively established.

### Possible interaction between heart rate and activity

An interaction term between heart rate and activity, specifically the ratio of minutes in fat burn heart rate zone and lightly active minutes was included to account for potential interaction between heart rate and activity, for example, elevated heart rate without accompanying activity could possibly be a proxy for physiological stress or autonomic dysfunction rather than exercise. While statistical significance has not been detected for this interaction term, it appears in top important features in XGBoost model for subcohorts such as OSA, MDD and ADHD (absolute mean SHAP values *>*0.1). Past studies have also extracted features from activity and heart rate data in parallel [14] where heart rate is stratified by activity zones before analysis. Using granular minute-level FitBit data to capture finer interaction between heart rate and activity could be a promising future research direction.

### Association between Deep and REM sleep with disease risk

Both higher minutes spent in deep and REM sleep were found to associate with lower disease risk in some subcohorts, consistent with literature. Deep (slow-wave) sleep is associated with decreased diabetes risk [15], decreased blood pressure, heart rate, and cardiovascular disease risk [16]. Some studies have also linked depression with decreased slow-wave sleep [17]. Decreased REM sleep has been linked to increased all-cause mortality [18]. Past literature has shown that higher variations in sleep duration, deep and REM sleep are associated with conditions such as atrial fibrillation, MDD and anxiety, etc [6]. While the only statistical significance detected among variation-related features is higher variation in REM sleep associating with higher disease risk in ADHD, STD and RMSSD of Deep and REM sleep were ranked as important features in some subcohorts in XGBoost models with absolute mean SHAP values *>*0.1.

### Difference between logistic regression coefficients and XGBoost feature importance

While statistical significance has been observed only for some activity and sleep features in logistic regression and no statistical significance has been observed for heart rate features, these features generally have higher absolute mean SHAP values in XGBoost models, whereas lifestyle features, especially drinking habits have low, sometimes near zero absolute mean SHAP values in XGBoost models even though these features are statistically significant in logistic regression. This is because Logisitic Regression coefficients and XGBoost feature importance measure different aspects of the data, the former measuring linear effect and the latter measuring contribution to prediction. A feature can have a large SHAP value and large contribution to prediction even if its main linear effect is weak; tree-based XGBoost models automatically captures threshold effects, local non-linear patterns and interaction with other features. The high SHAP values for wearable-derived features, for example, ratio of minutes in fat burn zone to lightly active minutes and RMSSD of sleep features, despite limited statistical significance, possibly indicate additional non-linear effects of these features not captured by logistic regression. In addition, XGBoost models achieving similar performance as logistic regression without relying on lifestyle features might indicate that wearable-derived features contain overlapping information as lifestyle features and could help to improve disease risk prediction even when lifestyle information is absent.

## Limitations

### Accuracy and noise of FitBit collected physiological data

A major limitation of this study is the reliance on FitBit summary statistics. As the underlying aggregation aggregation algorithms are proprietary, there is limited transparency and control over data quality and algorithms. Recent evaluations of FitBit devices indicate a tendency to overestimate time spent in higher-intensity activities [19]. For sleep metrics, meta-analyses have shown that FitBit devices have promising performance in sleep staging [20] with good accuracy, sensitivity but low specificity. For example, FitBit might tend to overestimate light sleep and underestimate wake after sleep onset [21]. Furthermore, reliability of such summary statistics varies across different FitBit versions, which were not differentiated in this study to preserve as many sample sizes as possible.

In addition, this study may be biased due to the use of a single cohort; validation in external studies with different commercial wearable devices in the future could help to mitigate the biases and assess the generalizability of these models and regression coefficients.

## Conclusion

Wearable-derived heart rate, activity and sleep features are associated with disease risks and improve disease classification capabilities beyond just demographic and lifestyle factors. This study is among the first comprehensive trans-diagnostic studies to examine this using large-scale real-world datasets. Simple machine learning models, including logistic regression and XGBoost, have achieved both good discrimination and calibration in disease classification, and could be explored as an auxiliary tool for patient disease risk alert. This study has also demonstrated that variations in sleep structures, as well as variations in and interactions between heart rate and activity, may be important for disease risk. Future studies could investigate more granular wearable-derived features to capture finer-scale variations and interactions in these physiological signals.

## Supporting Information

**S1 Table. Inclusion and Exclusion criteria based on ICD codes**

**S2 Table. List of medications excluded from Clean Control cohort**

**S3 Table. Descriptives of subcohorts**

**S4 Table. Comparison of control vs clean control cohort**

**S5 Table. Full comparison of model performance metrics**

**S6 Table. Full results for logistic regression coefficients and p-values**

**S7 Table. Mean Absolute SHAP values for XGBoost models calculated on test dataset**

## Data Availability

This study used data from the All of Us Research Program‘s Controlled Tier Dataset version 8, available to authorized users on the Researcher Workbench. All code used to generate the results in this study is publicly available at: https://github.com/xueyann/wearable-transdiagnostic-classification-allofus

## Acknowledgement

We gratefully acknowledge All of Us participants for their contributions, without whom this research would not have been possible. We also thank the National Institutes of Health’s All of Us Research Program for making available the participant data examined in this study.

## References

1. Stan IE, D’Auria D, Napoletano P. A systematic literature review of innovations, challenges, and future directions in telemonitoring and wearable health technologies. IEEE Journal of Biomedical and Health Informatics. 2025.

2. Lee S, Chu Y, Ryu J, Park YJ, Yang S, Koh SB. Artificial intelligence for detection of cardiovascular-related diseases from wearable devices: a systematic review and meta-analysis. Yonsei medical journal. 2022;63(Suppl):S93.

3. Kundrick J, Naniwadekar A, Singla V, Kancharla K, Bhonsale A, Voigt A, et al. Machine learning applied to wearable fitness tracker data and the risk of hospitalizations and cardiovascular events. American Journal of Preventive Cardiology. 2025:101006.

4. Jacobson NC, Weingarden H, Wilhelm S. Digital biomarkers of mood disorders and symptom change. NPJ digital medicine. 2019;2(1):3.

5. Jacobson NC, Feng B. Digital phenotyping of generalized anxiety disorder: using artificial intelligence to accurately predict symptom severity using wearable sensors in daily life. Translational Psychiatry. 2022;12(1):336.

6. Zheng NS, Annis J, Master H, Han L, Gleichauf K, Ching JH, et al. Sleep patterns and risk of chronic disease as measured by long-term monitoring with commercial wearable devices in the All of Us Research Program. Nature Medicine. 2024;30(9):2648–56.

7. Bailey CP, Dodd KW, McClain JJ, Seo I, Wheeler W, Wolff-Hughes DL. Fitbit physical activity and sleep data in the all of us research program: data exploration and processing considerations for research. Medicine and science in sports and exercise. 2025:10–1249.

8. Master H, Kouame A, Hollis H, Marginean K, Rodriguez K, Center DR, et al.. 2022Q4R9 v7 Data Characterization Report; 2024. Accessed January 14, 2026. https://support.researchallofus.org/hc/en-us/articles/14558858196628-2022Q4R9-v7-Data-Characterization-Report.

9. Research S. Through ‘All of Us’ program, Scripps Research launches wearable technology study to accelerate precision medicine; 2021. Accessed January 14, 2026. https://www.scripps.edu/news-and-events/press-room/2021/20210224-aou-fitbit-study.html.

10. All of Us Research Program. Participant Privacy Protections; 2025. Updated February 03, 2025. Accessed January 14, 2026. https://support.researchallofus.org/hc/en-us/articles/4552681983764-Participant-Privacy-Protections.

11. Carroll RJ, Bastarache L, Denny JC. R PheWAS: data analysis and plotting tools for phenome-wide association studies in the R environment. Bioinformatics. 2014;30(16):2375–6.

12. Brier GW. Verification of forecasts expressed in terms of probability. Monthly Weather Review. 1950;78(1):1–3.

13. Murphy AH. A new vector partition of the probability score. Journal of Applied Meteorology. 1973;12(4):595–600.

14. Zhou W, Chan YE, Foo CS, Zhang J, Teo JX, Davila S, et al. High-resolution digital phenotypes from consumer wearables and their applications in machine learning of cardiometabolic risk markers: Cohort Study. Journal of Medical Internet Research. 2022;24(7):e34669.

15. Tasali E, Leproult R, Ehrmann DA, Van Cauter E. Slow-wave sleep and the risk of type 2 diabetes in humans. Proceedings of the National Academy of Sciences. 2008;105(3):1044–9.

16. Korostovtseva L, Bochkarev M, Sviryaev Y. Sleep and cardiovascular risk. Sleep Medicine Clinics. 2021;16(3):485–97.

17. Cheng P, Goldschmied J, Deldin P, Hoffmann R, Armitage R. The role of fast and slow EEG activity during sleep in males and females with major depressive disorder. Psychophysiology. 2015;52(10):1375–81.

18. Zhang J, Jin X, Li R, Gao Y, Li J, Wang G. Influence of rapid eye movement sleep on all-cause mortality: a community-based cohort study. Aging (Albany NY). 2019;11(5):1580.

19. Feehan LM, Geldman J, Sayre EC, Park C, Ezzat AM, Yoo JY, et al. Accuracy of Fitbit devices: systematic review and narrative syntheses of quantitative data. JMIR mHealth and uHealth. 2018;6(8):e10527.

20. Haghayegh S, Khoshnevis S, Smolensky MH, Diller KR, Castriotta RJ. Accuracy of wristband Fitbit models in assessing sleep: systematic review and meta-analysis. Journal of medical Internet research. 2019;21(11):e16273.

21. Lim SE, Kim HS, Lee SW, Bae KH, Baek YH. Validation of fitbit inspire 2TM against polysomnography in adults considering adaptation for use. Nature and Science of Sleep. 2023:59–67.

